# Quality of Systematic Reviews in Dental Journals Published from India: A Methodological Review

**DOI:** 10.1101/2023.06.06.23291040

**Authors:** Rohan Thomas Mathew, Anu Babu

## Abstract

**Background:** Evidence-based medicine (EBM) is a crucial aspect of modern medical practice, emphasizing the use of the best available evidence to inform clinical decision-making. Systematic reviews (SRs) play a key role in EBM by integrating and evaluating findings from multiple studies. However, the methodological quality of SRs can vary, and assessing their quality is essential for accurate interpretation and application of the findings.

**Methods:** This study aims to analyze the methodological quality of SRs published in dentistry journals from India between 2015 and 2020. A comprehensive search was conducted, resulting in the inclusion of 130 SRs from 21 journals.

**Results:** The findings reveal several discrepancies in the methodological quality of the included SRs. Only a small percentage of SRs were registered in the PROSPERO registry, and adherence to PRISMA guidelines was limited. The majority of SRs did not assess scientific quality, such as risk of bias and publication bias, adequately. Furthermore, the study highlights a lack of awareness and support for conducting high-quality SRs, including limited funding and insufficient utilization of standardized guidelines.

**Conclusion:** The authors suggest that increasing awareness among researchers, editors, and funding agencies, as well as adopting standardized guidelines and protocols, can improve the quality and quantity of SRs produced in India. Overall, this study emphasizes the need to enhance the methodological rigor of SRs in dentistry to ensure reliable evidence for clinical decision-making.

## Introduction

Evidence based medicine (EBM) is one of the most essential aspects of modern medical practice. It is considered one of the most important developments in medicine on par with antibiotics and anaesthesia. [1]. The first principle of EBM is that not all evidence is created equal and that the practice of medicine should be based on the best available evidence. The second principle is that the pursuit of the best available evidence is accomplished by evaluating the totality of evidence and not the ones that favour a particular claim. The third principle is that the clinical decision should give importance to the patient’s interest equally. [1]

A systematic review (SR) is a means to identify, appraise and integrate the findings of studies on a specific topic using a systematic approach based on previously established criteria employing methods that minimise bias through a rigorous and replicable methodology. [2, 3] Presently, this is considered as the gold standard for decision-making, creating guidelines by clinicians and policymakers and as the foundation for EBM. [3] It is also important for preventing duplication of research and for creating new avenues of trials.

The number of scientific publications occurring today is staggering. MEDLINE alone contains an overwhelming number of 22 million indexed citations from over 5600 journals which publishes 75 RCTs and 11 SRs daily. [1] The situation is no different in dentistry or oral health. There were 1188 SRs published in dentistry according to one study. [3] Few studies have reviewed the SRs published in dentistry and its various specialities. [2,3,4,5,6]

The validity and reliability of the conclusions arrived in an SR depends on the process. Appropriate methodology is essential for an accurate interpretation and application of these findings in clinical practice. Hence it is essential to assess the methodological quality of SRs. [5]

But currently, there are no studies which have studied the quality of SRs coming from a particular country. The number of dental colleges have increased in India in the last decade or so. The number of dentistry-related journals has increased in India in the last few years and many of these journals are regularly publishing systematic reviews. It is in this context that the authors decided to study the SRs which are published in India. We intend to identify these SRs and analyze them for their methodological quality.

## Methodology

The aim of the study was to do a methodological study of the SRs in dentistry published from India from 2015 to 2020. Since this was a methodological study the protocol was neither published nor registered. A comprehensive search was performed in the Medline, NLM catalogue and google scholar to identify journals published from India. A hand search was also performed to identify specialty and other general dentistry journals.

Both indexed and non – indexed journals are included in the study. Two authors went through the journals individually and identified all relevant titles published during the time period. The search was limited to studies published between January 2015 and December 2020.

### Study selection and eligibility criteria

The titles and abstracts of studies under the heading reviews from the selected journals were screened first. The full texts were retrieved for the potentially relevant studies. All systematic reviews were included. Mini reviews, narrative reviews and case series with literature reviews were excluded. Methodological studies were also excluded. We also excluded articles which were not available in full text.

### Data extraction

A data extraction form was created in MS Excel. A pilot data extraction using 5 random SRs were done. All the authors reviewed this and discussed further on the data and details to be included. A rule book was also created for the ease of entering data based on the discussion. Subsequently a final data extraction tool was made, which was followed for the study. Two authors reviewed the studies and decided on the titles to be included. Following which one author extracted the data from each SR. and another author independently checked all data. The data specific to the journal as well as the paper were collected. It included general details of the journal such as name, type of specialty, indexed, if so which index and whether the journal is predatory or not. Since there are no clear guidelines of defining predatory behavior, the journals are rated as less likely, likely or more likely to be predatory. Presence in the Beall’s list of predatory journals / publishers will make it more likely. If indexed, then it is less likely. If there is only a short time duration from date of submission to date of acceptance, without adequate time for a good review then also it is considered to be predatory. The authors also used AMSTAR (A Measurement Tool to Assess systematic Reviews) instrument to assess the methodological quality of the reviews included for the study. This tool uses 16 items to identify if the review under question is critically low, low, moderate or high quality. [13]

The data extraction form also included details about the paper such as the number of authors, author composition; whether they are from Indian or foreign institutes, PROSPERO registration, reporting of search strategy, whether PRISMA guidelines are followed, number of databases, among which how many are academic and how many are grey literature, details of review process, risk of bias appraisal, quality of evidence, details of meta-analysis if it is done, software used and details of funding if received. The author only checked whether these are reported or not. The validity or accuracy of the parameters are not checked. Discrepancies were discussed and solved by consensus or by adjudication by the third author.

### Data Analysis

The data obtained is tabulated, and descriptive statistics are used to analyse the data.

## Results

The total number of systematic reviews included in the study are 130. They are from 21 journals covering all specialties, general dentistry and a few interdisciplinary journals. 83% of the reviews have 3 or more than 3 authors. General dentistry journals have published 49 reviews which is the maximum number of reviews among all the specialties. Close to 85% of the reviews are published in indexed journals. Around 71 % of the reviews were published in journals which are less likely to be predatory. Among the reviews included in the study 38 are published in 2019 which is the highest.

Only 15% of the reviews have been registered in PROSPERO registry. 50% of the reviews have followed PRISMA guidelines. 83% of the reviews have mentioned the search strategy while the remaining have not made any mention of the search strategy used. 86% of the reviews have mentioned inclusion and exclusion criteria. 65 % of the reviews have searched only in 3 or less academic databases.

63 % of the reviews says they have done double screening of the articles and only 34% of the reviews have done adjudication in case of disagreement by the independent reviewers.

55% of the studies have done risk bias appraisal while only 4% studies have done assessment of publication bias. Of the reviews we have taken for this study 25% have done meta-analysis. 8% of the studies have performed synthesis of results. Out of 130, 116 reviews mentioned whether funding is received or not. Only 4 reviews have received funding. The AMSTAR tool scores the reviews as critically low, low, moderate or high. The tool showed that 65% of the reviews are of critically low quality. Among the remaining studies 18% are of low quality, 16% are moderate quality and 1% is high quality.

**Figure 1:**
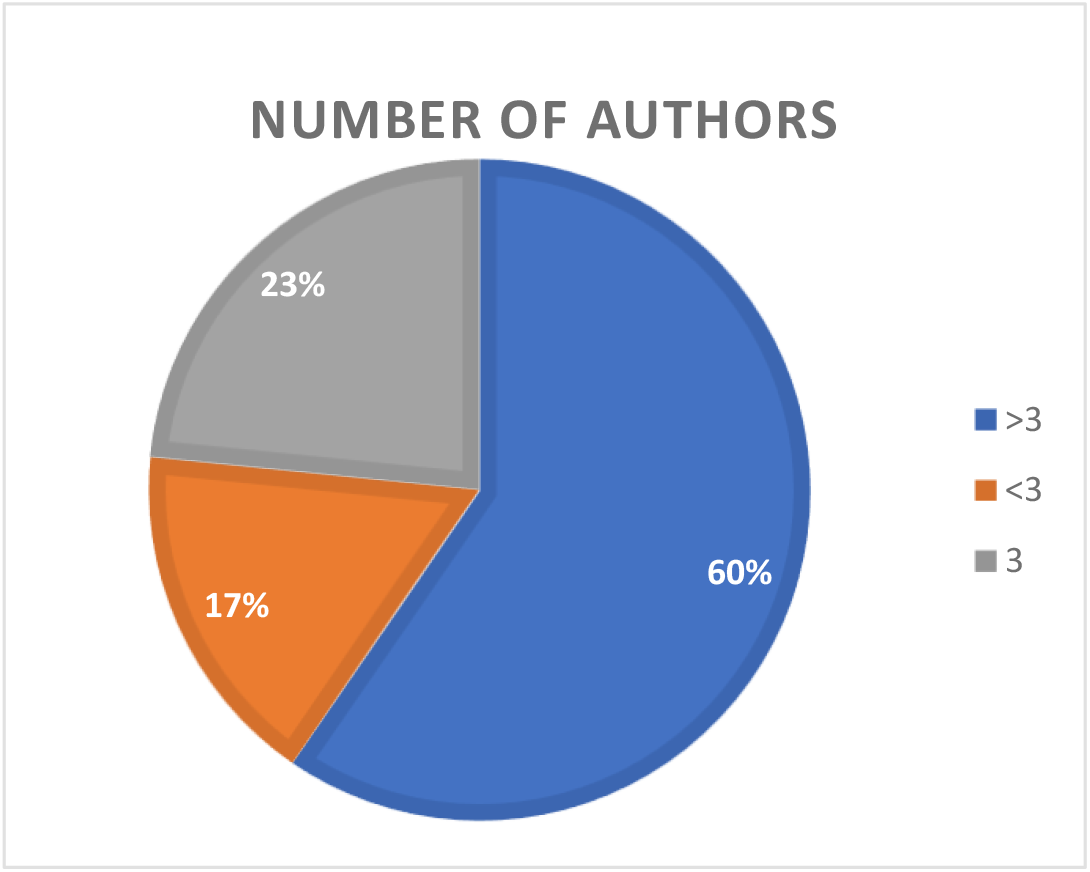
Number of authors

**Figure 2:**
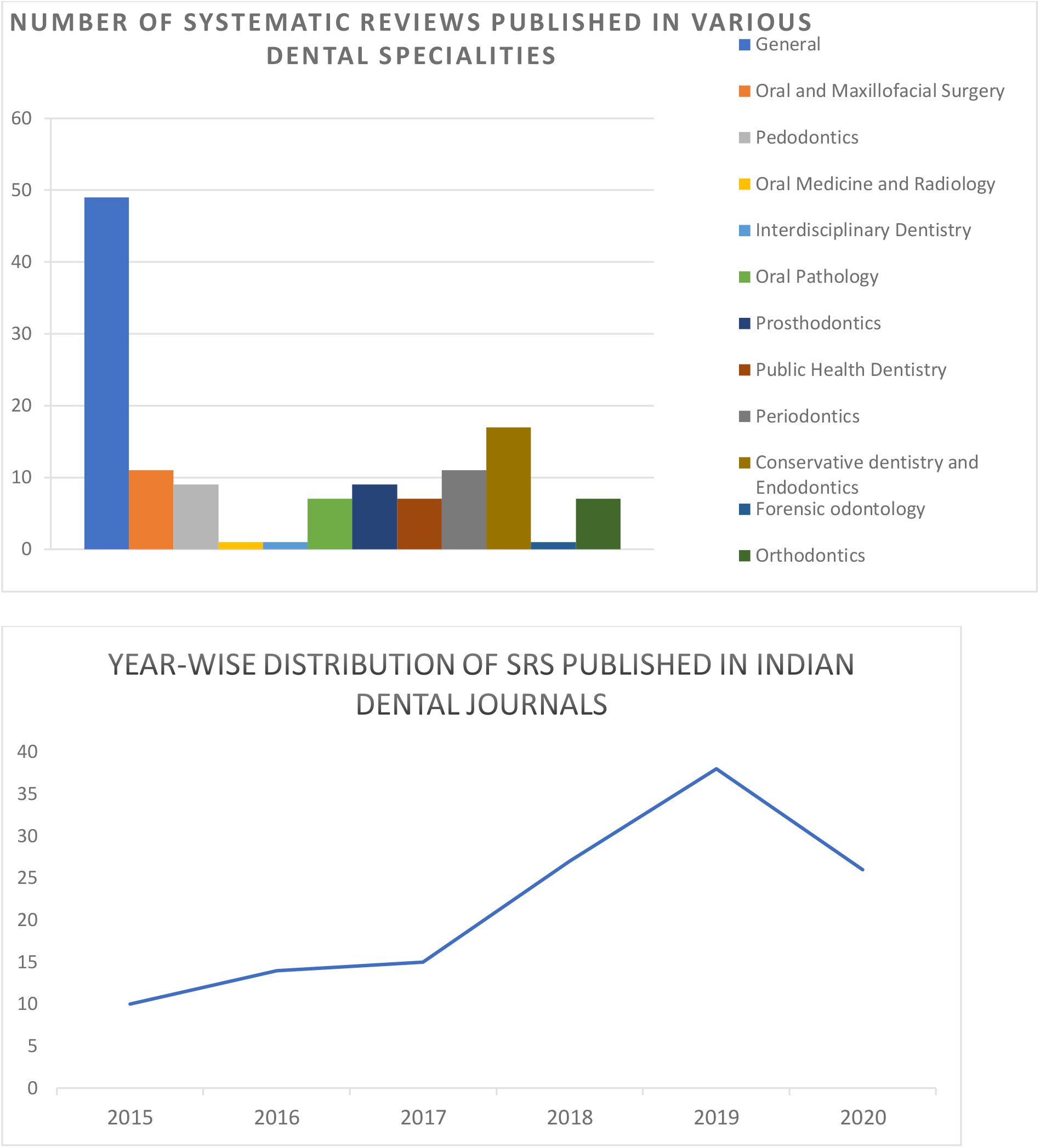
Number of SRs in various

## Discussion

In this age of information explosion, considering the inherent human brain limitations in processing evidence application of EBM principles to identify the high-quality research is mandatory. EBM is the cornerstone of modern medical practice. It consists of clinical guidelines which are based on SRs of well conducted RCTs. [1] The growing number of trials and studies coming out in dentistry has led to new challenges for an individual practitioner to take clinical decisions.

The total number of SRs we identified are 130 which is a modest number when compared to the number of dentists and dental schools that are there in India. A similar study was conducted by Bassani et al who studied all the SRs in Dentistry in 2017 from PubMed. The highest number of SRs from any country was from Brazil which had 117 SRs in 2017 alone. The same year there was only 15 SRs from India. The reason they suggest is that in Brazil, the promotions and appointments in universities are based on the publications. [2] The same has been applied in India but there was no increase in the publication of SRs. The journal editors should invite and support more systematic reviews. In addition, there is overall lack of funding for the publication of SRs.

Although an increase in the number of SRs may not reflect an improvement in the methodological quality. One of the principles of EBM is the evaluation of the totality of evidence. Close to 17% of the SRs had less than 3 authors. 63 % of the reviews have done double screening which only 34% have had adjudication by an independent reviewer in case of disagreement. 65% of the SRs have searched in 3 or less academic databases. Having at least two reviewers to select relevant studies and extract data in duplicate reduces the potential biases and helps reduce accidental exclusion of relevant data which may lead to distorted conclusions. Failure to search multiple databases may also lead to the exclusion of relevant studies which can produce biased results. [3]

The assessment of scientific quality of studies is important to understand whether the recommendations from the SR are reliable to change existing practice. While 55% of the reviews have done risk of bias appraisal only 4% of the reviews have done publication bias. In a methodological review of SRs in Endodontic specialty, the same was observed. The majority of the reviews in that study also did not assess the scientific qualities of the studies included. [5]

The first principle of EBM is that not all evidence is created equal. If, during the course of conducting an SR, the reviewers do not assess the quality of evidence included, then the conclusions are on shaky grounds.

The PRISMA statement, Cochrane Collaboration, Joanne Briggs Institute (JBI) have helped the reviewers standardize the process of conducting a review. They have put down guidelines and manuals for the authors to follow for the proper conduct of SRs. [7,8] We have found that only 50% of the reviews have mentioned that they have followed PRISMA guidelines. This shows a widespread lack of awareness about how to perform an SR. Bassani et al in a similar study have also said that using PRISMA guidelines is suboptimal in dentistry. Even in the ones that report the use of PRISMA statement do so without having a deeper knowledge on it. [2] This also points to the lack of awareness about such guidelines among the reviewers and editors as well.

Another important aspect of a systematic review is protocol preparation and registration of the said protocol in PROSPERO registry which is international database of prospectively registered systematic reviews. [9] This avoids unintended duplicity of the work. It also helps reduced the reporting bias as the reader or the editor can compare the final review and the protocol which was planned. Our study found out that only 15% of the reviews are registered in PROSPERO. This is quite low, and the onus is on the editors to insist on PROSPERO registration for the protocol.

When compared to other methodological studies about SRs in dentistry we found that a significantly high number of SRs are published in general dentistry. The reviews published in the general dentistry journals comprise of topics from all specialties. The specialty journals should encourage more reviews to be published in their respective specialties.

The funding agencies should also encourage to grant more funds to conducting SR. In our study only 4 reviews received funding out of which 3 are by authors from outside India. The lack of support for the authors to conduct a thorough review often worsens the quality of evidence created. Professional organisations and specialty organisations are in a much better place to identify gaps in the present evidence and provide support for conducting an SR.

The number of reviews publishing from India are gradually increasing which is a good sign but when compared to other countries, the number is abysmally low. [2] The overall quality needs to be improved and there are enough and more material available from Cochrane collaboration and JBI on how to conduct a good systematic review. [10]

The authors used AMSTAR instrument also to assess the quality of the reviews as this is a standardized and widely accepted tools to measure an SR. The tool revealed that 65% of the SRs which were studied are of critically low quality. Only 1 SR was of high quality. This correlates with the authors’ assessment that a lot is left to be desired.

Our study was aimed to analyze the methodological qualities of SRs published in journals from India, due to which a comprehensive search across databases was not effective. The authors then hand searched journals related to dentistry from India to identify the SRs. This is a limitation of the study as some of the reviews may not have been included in the study. The authors also did not validate what was reported in the study to what was done in the study. For example, the authors checked if the search strategy is given or not; the validity or relevance of the search string with respect to the research question is not checked. This is a drawback of our study. The authors have done predominantly hand-searching when it came to identifying journals and the published reviews hence some journals may be missed.

## Conclusion

This methodological review has found that there are several discrepancies in the reviews published from India. This could lead to biased and possibly incorrect results. Inclusion of more databases, including a greater number of authors, assessing the quality of evidence and following standard guidelines or checklists can significantly increase the quality of the evidence created. Increased awareness about the nuances of systematic reviews among the editors is also needed. India has 315 dental colleges, and 269 of them are running post-graduation programs presently, which is one of the highest in the world. [11] Adequate support from the funding agencies can immensely help to increase the quantity and quality of evidence created from India.

## Data Availability

All data produced in the present study are available upon reasonable request to the authors

## Acknowledgement

The authors would like to acknowledge the contributions of Dr.Denny John in the conception and formulation of the study.

